# Quantifying and visualising heterogeneity in cumulative adverse childhood experiences scores

**DOI:** 10.64898/2025.12.09.25341822

**Authors:** Bruna Cardone Pedroso Elorza, Alicia Matijasevich, Maria Fernanda Tourinho Peres, Andreas Bauer

## Abstract

**IMPORTANCE:** Adverse childhood experience (ACE) scores mask substantial heterogeneity within the 2-, 3-, and ≥4-ACE exposure categories. The magnitude of this heterogeneity has not been sufficiently quantified.

**OBJECTIVE:** To quantify and visualise the heterogeneity of ACE scores.

**DESIGN:** Cross-sectional analysis of the 2023 National Survey of Children’s Health, an address-based, nationally representative survey.

**SETTING:** US households.

**PARTICIPANTS:** A national sample of 385,000 addresses was drawn. Between 23 June 2023 and 19 January 2024, caregivers of children aged 0–17 years completed screening and topical questionnaires; 55,162 of 70,187 eligible households completed the topical questionnaire.

**EXPOSURES:** Lifetime exposure to 10 ACEs, assessed via caregiver report: economic hardship; parental divorce/separation; household mental illness; household substance use; parental incarceration; domestic violence; racial discrimination; neighbourhood violence; health- or disability-based discrimination; and parental death.

**MAIN OUTCOMES AND MEASURES:** Heterogeneity of ACE co-occurrence within the 2-, 3-, and ≥4-ACE exposure categories, quantified using combinatorial coverage (CC; proportion of possible co-occurrence patterns observed) and visualised with an UpSet plot of item combinations with unweighted *n* ≥ 30.

**RESULTS:** Analyses used complete ACE data (*N* = 51,468; 93.3% of the total sample); co-occurrence analyses were restricted to children with ≥2 ACEs (*n* = 7,897). Across the 2-, 3-, and ≥4-ACE categories, 582 of 1,013 possible combinations were observed: all 45/45 patterns for 2 ACEs (CC_2_ = 1.00), 110/120 for 3 ACEs (CC_3_ = 0.917) and 427/848 for ≥4 ACEs (CC≥4 = 0.504). Of the 582 distinct combinations, 50 met the display threshold, accounting for 5,204 (66%) of the 7,897 children with ≥2 ACEs: 23, 15 and 12 for 2, 3 and ≥4 ACEs, respectively. Certain ACEs recurred disproportionately among these intersections: parental divorce/separation appeared in 34 of 50 patterns, household substance use in 23 and household mental illness in 20.

**CONCLUSIONS AND RELEVANCE:** In this nationally representative sample of US children, ACE scores masked substantial heterogeneity in exposure patterns within the 2-, 3-, and ≥4-ACE categories. Simple heterogeneity diagnostics, such as combinatorial coverage and UpSet plots, can make this heterogeneity explicit and may prompt pattern-specific analyses of outcomes.

**Key Points:** *Question:* How heterogeneous are cumulative adverse childhood experience (ACE) scores, and how can this heterogeneity be quantified and visualised?

*Findings:* In a nationally representative cross-sectional survey of 51,468 US children, 582 of 1,013 theoretically possible ACE combinations among those with ≥2 ACEs were observed: all 45/45 patterns for 2 ACEs, 110/120 for 3 ACEs and 427/848 for ≥4 ACEs. When visualised with an UpSet plot, some individual ACEs recurred disproportionately among the 50 most common co-occurrence patterns, which together accounted for 5,204 (66%) of the 7,897 children with ≥2 ACEs.

*Meaning:* ACE scores mask substantial heterogeneity within the 2-, 3- and ≥4-ACE exposure categories, risking misleading inferences about which children are at highest risk; this heterogeneity can be made explicit using a simple metric and visualisation technique.

---

Adverse childhood experiences (ACEs) encompass childhood abuse, neglect, and household dysfunction.^1^ They are highly prevalent – around six in ten adults worldwide report at least one ACE^2^ – and are associated with impairments across multiple domains of health and functioning, including physiological regulation, neurocognitive development, chronic disease, psychopathology, maladaptive behavior, and involvement with the criminal justice system.^3^ ACEs also impose substantial economic burdens, exceeding half a trillion US dollars annually.^4^

The ACE framework was originally conceptualised as a *cumulative risk approach*,^1^ which remains widely used in contemporary research.^a^ This approach sums the number of ACEs and categorises individuals into exposure groups of 1, 2, 3, or ≥4 ACEs. While the resulting ‘ACE score’ has several strengths (for a discussion, see Lacey & Minnis, 2020),^5^ it captures only quantitative (count-based) rather than qualitative (type-specific) variation. Consequently, individuals with the same ACE count are treated as a homogenous group, despite likely having markedly different exposure profiles. It also implicitly assumes equal weighting of items – an assumption both theoretically questionable and often empirically unsupported.

The issue of heterogeneity in cumulative risk scores was investigated by Barger and Oláis,^6^ who used Behavioral Risk Factor Surveillance System (BRFSS) 2019–2020 data to examine associations between the five most common ACE combinations within each exposure category and two outcomes: current smoking and lifetime depression. Their analysis revealed substantial heterogeneity in effect sizes. For example, among individuals with three ACEs who reported both physical and verbal abuse, those who also reported parental divorce had an incidence rate ratio of 1.82 for lifetime depression, whereas those who instead reported household mental illness had a ratio of 4.12 – that is, replacing a single item more than doubled the risk. Furthermore, some effect sizes for two-ACE combinations

(e.g., verbal abuse and household mental illness) exceeded several three- or four-ACE combinations (e.g., verbal abuse, parental divorce, and household substance use). Taken together, these findings suggest that the *types* of adverse events, not just their *numbers*, are crucial.^7^ They also highlight the importance of additive synergistic interactions, in which certain ACE pairs contribute more than the sum of their individual effects,^8^ and of possible non-linear (saturation) effects for some outcomes.^9^ While this work represents a major advance, it also has limitations: among more than 9,000 participants with three ACEs, over 6,000 were grouped into an undifferentiated “all other triple ACEs” category and excluded from detailed analysis; and identifying the most common item combinations required additional coding steps.

With 2^*k*^ possible combinations (1,024 for the original 10-item ACE questionnaire), interpreting heterogeneity in cumulative risk scores is inherently challenging. Visualisation techniques can assist in uncovering complex patterns of co-occurrence. The UpSet plot was specifically designed to address this ‘combinatorial explosion’: it visualises set intersections by encoding them as binary patterns in a matrix (present/absent), with accompanying bar charts to show intersection sizes.^10^ While originally developed and tested for areas such as cancer biology and economics,^10^ applications have been discussed across various domains.^11,12^ However, to our knowledge, no published studies have yet applied the UpSet plot to ACE research.

To address this gap, the objectives of the current study are twofold: (1) to quantify and visualise heterogeneity in cumulative ACE scores in a large, representative sample of US children – both overall and within each exposure category – and (2) to introduce the UpSet plot as a visualisation tool for the ACE framework.

## METHODS

### Sample

We used data from the 2023 National Survey of Children’s Health (NSCH),^b^ an address-based, nationally representative, cross-sectional survey of US households with children aged 0–17 years that measures health and well-being across children, families, and communities. In 2023, 385,000 national addresses were sampled; 122,000 screeners were completed, 70,187 households were eligible for the topical questionnaire, and 55,162 interviews were completed (overall weighted response rate 35.8%). All analyses applied child-level survey weights, allowing generalisation to state and national resident child populations; a non-response bias analysis found no strong or consistent evidence of bias after weighting.

### Measures

Lifetime exposure to ten ACEs was assessed via caregiver report. The items were: economic hardship; parental divorce/separation; household mental illness; household substance use; parental incarceration; domestic violence; racial discrimination; neighbourhood violence; health- or disability-based discrimination; and parental death. All items were binary (yes/no) except economic hardship, asked on a four-point scale. Following Lu,^13^ we coded ‘never’ or ‘rarely’ = 0 and ‘somewhat often’ or ‘very often’ = 1; an alternative threshold (‘never’ = 0 and ‘rarely’ or more = 1) was tested in sensitivity analyses. Full item wording and original response options are provided in eTable 1.

### Statistical analysis

Analyses proceeded in four steps. First, we recoded all ACE items as binary (0□=□no, 1□=□yes) and defined ACE-score exposure categories of 0, 1, 2, 3, or ≥4 ACEs; co-occurrence analyses focused on 2, 3, and ≥4 (children with ≥2 ACEs; *n* = 7,897). Second, we defined combinatorial coverage (CC) as the proportion of possible co-occurrence patterns that were observed within each exposure category. Formally, for exactly *k* ACEs, 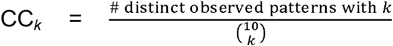(for *k* = 2, 3); for ≥4 ACEs,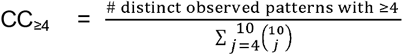 . CC ranges from 0 to 1 (0-100%), with higher values indicating greater coverage. Third, we generated an UpSet plot (restricted to children with ≥2 ACEs), highlighting exposure categories involving 2, 3, and ≥4 ACEs using distinct colours. The presentation of co-occurrence patterns was limited to those with unweighted *n□*≥ 30, in line with NSCH guidance. Because weighting changes counts but not the underlying co-occurrence structure, figures present unweighted intersection sizes. To provide population context, we report survey-weighted estimates for the most common intersections, which generalise to the US resident child population. Fourth, we conducted a sensitivity analysis applying a less stringent threshold for economic hardship. We restricted the analytic sample to complete cases on all ten ACEs because item-level missingness was low (<5% per ACE; see eTable 2). For illustrative purposes, we also visualised missing-data patterns using an UpSet variant to demonstrate additional applications and to investigate whether specific ACE combinations were missing. Prevalence rates of individual ACEs have been reported elsewhere and are not discussed further.^14^ All analyses were conducted in RStudio (2023.12.0) with the R packages: ggplot2 3.5.1, ComplexUpset 1.3.6, and naniar 1.1.0. All analysis code will be made openly available in an online repository, linked from the preprint (doi: XXX) and from the final published article.

## RESULTS

Analyses used complete cases for all 10 ACE items (*N*□=□51,468; 93.3% of the original sample). An UpSet⍰style plot of item⍰level missingness showed no evidence of clustering in specific ACE combinations; excluded cases typically had missingness across all ACEs or on individual items (see eFigure 1).

To quantify heterogeneity, we compared the number of distinct observed co-occurrence patterns with the number of possible combinations in each ACE exposure category (see Figure 1). Among those with exactly two ACEs, all 45 possible combinations were observed (CC_2_□=□1.00); among those with exactly three ACEs, 110 of 120 were observed (CC_3_□=□0.917); and among those with ≥4 ACEs, 427 of 848 were observed (CC_≥4_□=□0.504). Thus, across the three ACE exposure categories, 582 of 1,013 possible combinations were observed.

**Figure 1.**
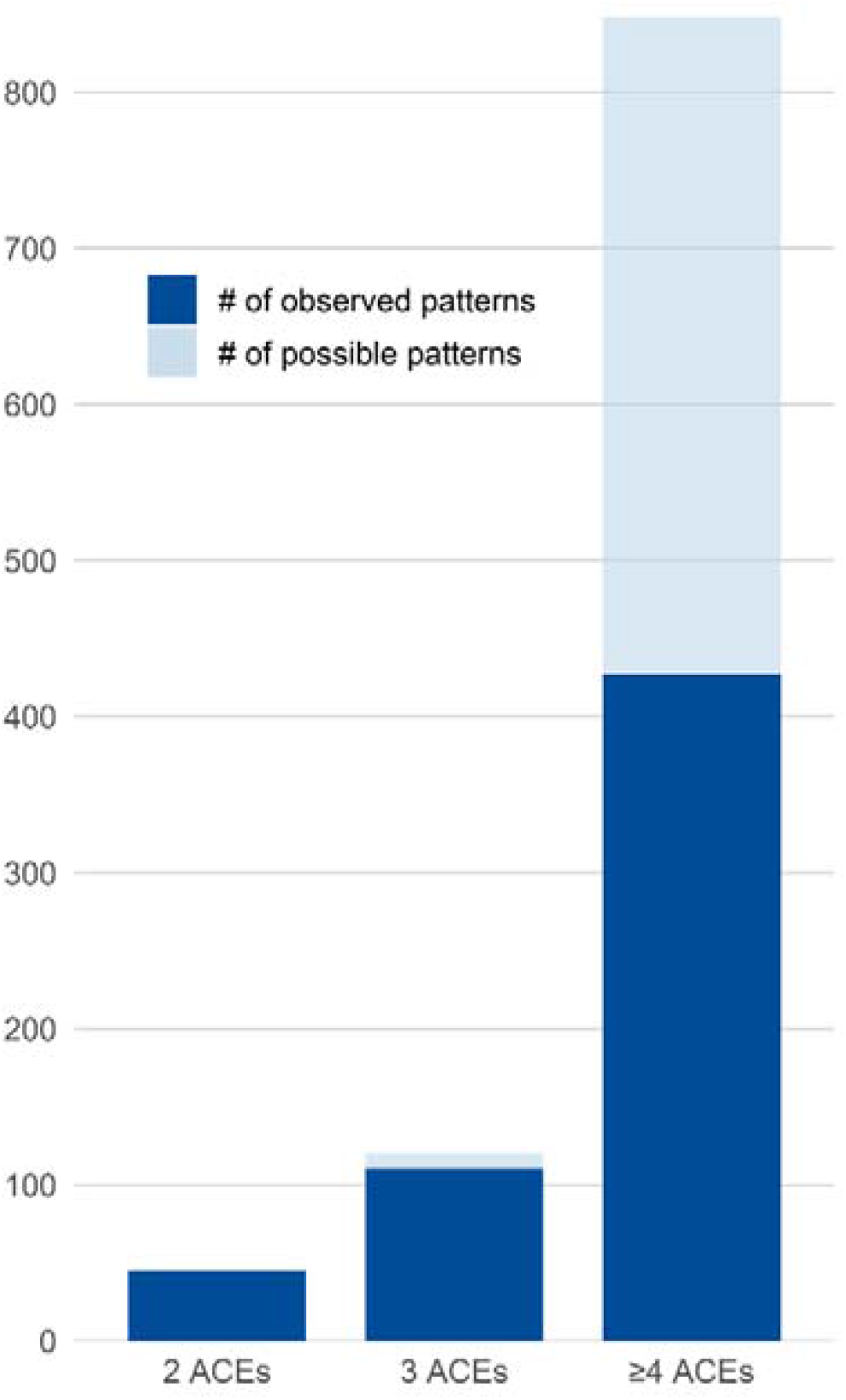
Observed vs. possible ACE item combinations, by exposure category ***Note***. Based on complete cases for all ACE items (*N* = 51,468); figure shows children with ≥2 ACEs (*n*□=□7,897). Bars show the total number of possible item combinations for each exposure category (2 ACEs: 45; 3 ACEs: 120; ≥4 ACEs: 848). The dark segment indicates observed combinations in the sample; the light segment indicates possible but unobserved combinations.

Of the 582 distinct ACE combinations, 50 met the display threshold of unweighted *n* ≥ 30, accounting for 5,204 (66%) of the 7,897 children with ≥2 ACEs, while the remaining 2,693 children (34%) were spread across 532 rarer combinations below the display threshold. Specifically, 23, 15 and 12 of the displayed patterns involved 2, 3, and ≥4 ACEs, respectively (see Figure 2). Unweighted group sizes for these displayed patterns ranged from 31 to 707 for 2 ACEs, 30 to 213 for 3 ACEs and 30 to 83 for ≥4 ACEs. Across exposure categories, certain ACEs recurred disproportionately among intersections: parental divorce/separation appeared in 34 of the 50 patterns, household substance use in 23 and household mental illness in 20.

**Figure 2.**
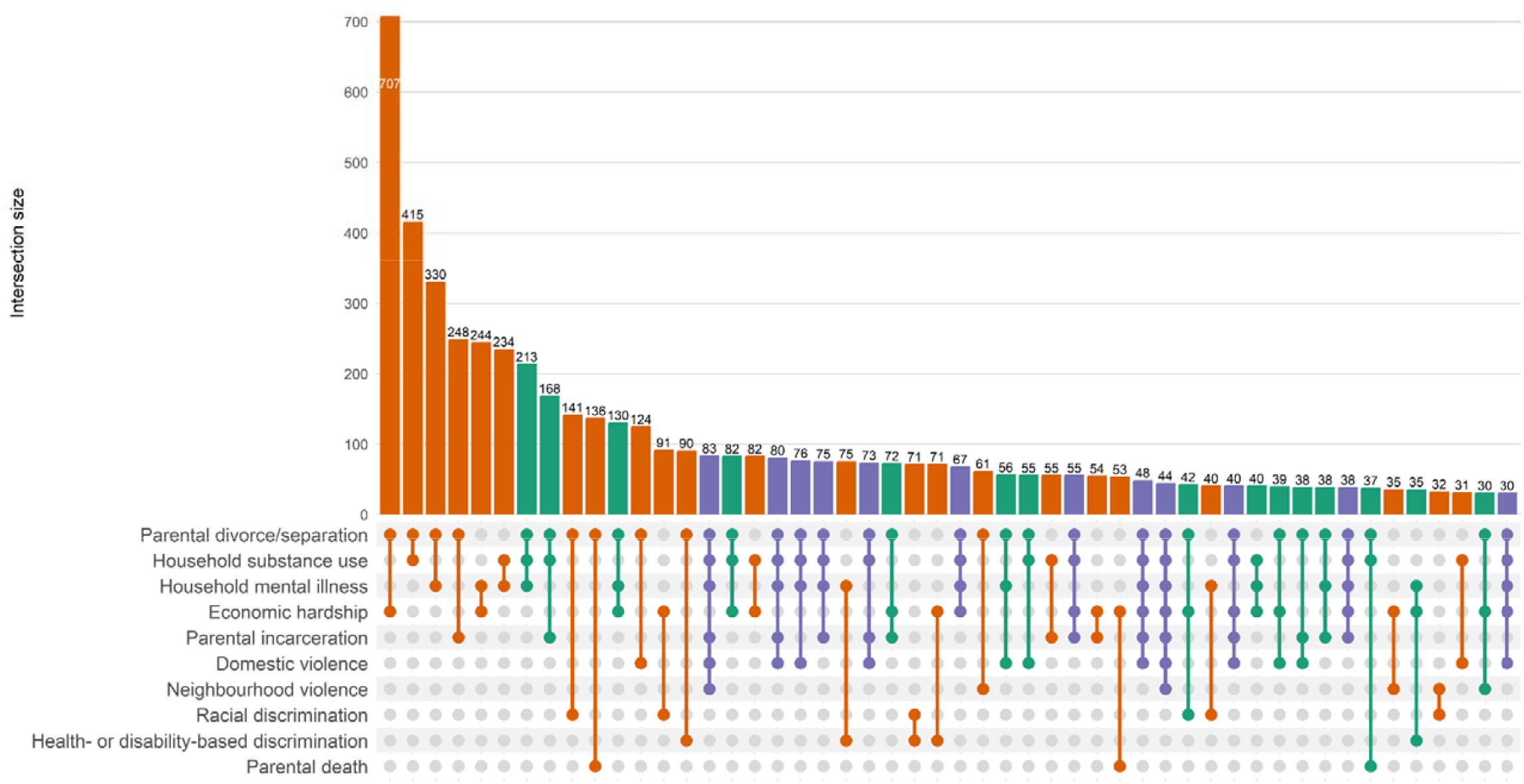
Observed ACE item combinations, coloured by exposure category ***Note***. Based on complete cases for all ACE items (*N* = 51,468); figure shows children with ≥2 ACEs (*n*□=□7,897). Columns show observed intersections with unweighted *n□*≥ 30, ordered in descending order by intersection size. The dot matrix shows which ACE items co□occur in each intersection; top bars show unweighted counts. Colours indicate exposure count: 2 ACEs (orange), 3 ACEs (green), ≥4 ACEs (purple).

For exactly two ACEs, the most common pairings were parental divorce/separation with economic hardship (weighted *n*□=□1,280,888), household substance use (*n*□=□425,011), or household mental illness (*n*□=□359,339). For exactly three ACEs, the most frequent combinations were: parental divorce/separation with household substance use and household mental illness (*n*□=□247,752); parental divorce/separation with household substance use and parental incarceration (*n*□=□209,371); and parental divorce/separation with household mental illness and economic hardship (*n*□=□189,956). For ≥4 ACEs, the most common combinations comprised the triad of parental divorce/separation, household substance use, and household mental illness together with: parental incarceration, domestic violence, and neighbourhood violence (*n*□=□118,211); parental incarceration and domestic violence (*n*□=□100,008); or domestic violence (*n*□=□98,122).

In a sensitivity analysis using a less stringent threshold for economic hardship, coverage remained 45/45 for 2 ACEs (CC_2_□=□1.00) but declined for three ACEs (92/120; CC_3_□=□0.767) and ≥4 ACEs (414/848; CC_≥4_□=□0.488) (see eFigure 2). Under this threshold, economic hardship became the most dominant ACE (35 times), especially in the 2-ACE pairing with parental divorce/separation, which outnumbered all other combinations by a wide margin (see eFigure 3).

## DISCUSSION

Using a nationally representative US sample, we paired a simple metric – combinatorial coverage (CC) – with a novel visualisation technique – UpSet plot – to characterise heterogeneity in cumulative ACE scores. We observed a wide variety of co-occurrence patterns within the 2-, 3-, and ≥4-ACE exposure categories, indicating that identical ACE counts can reflect markedly different exposure profiles and, consequently, potentially different levels of risk.^6^

To quantify heterogeneity, we calculated CC: the proportion of theoretically possible ACE colzloccurrence patterns observed within each exposure category. In total, 582 of 1,013 possible combinations were realised across categories. CC is simple to compute and reveals ‘pattern richness’, but it is dependent on sample size and on the items used; lowerl⍰lcount categories tend to yield higher values because their denominators are smaller (e.g., 45 for 2 ACEs vs 848 for ≥4 ACEs). Accordingly, CC is most informative within a given count category and when comparing subgroups or datasets that use the same ACE items (e.g., between male and female children or across 2016-2023 NSCH). Comparisons across exposure categories (e.g., 2 vs. 3 vs. ≥4 ACEs) should be made cautiously and used primarily to contextualise heterogeneity rather than rank levels of heterogeneity.

To visualise heterogeneity, we used an UpSet plot, which encodes set intersections as a binary (present/absent) matrix with accompanying bars for intersection sizes. Limiting display to intersections with unweighted *n*□≥ 30 yielded 50 distinct exposure profiles; among these, parental divorce/separation featured most frequently. UpSet plots are most interpretable when the number of displayed sets and intersections is moderate; Lex et al.^10^ note a practical limit of 40-50 sets, and readability similarly declines as the number of displayed intersections grow. Because only 50 of 582 observed combinations met the *n*□≥ 30 threshold, accounting for 66% of children with ≥2 ACEs, the figure should be read as a summary of higher-prevalence item combinations rather than an exhaustive catalogue. Thresholds should follow dataset-specific disclosure rules, which may vary across samples. When >50 intersections exceed the threshold, creating separate figures by

ACE-count category improves readability. UpSet also supports sorting, grouping by set degree, and annotation of intersection-specific statistics, which can reveal structure not apparent in bar charts. We generated plots using ComplexUpset, which integrates with ggplot2 and allows extensive customisation; similar figures can also be produced with UpSetR or ggupset.

These findings have several implications for research. First, count-based ACE scores can obscure substantial within-category heterogeneity, potentially biasing effect estimates and limiting their utility for identifying children at highest risk.^5^ Second, approaches that reduce ACEs to a small number of latent classes (e.g., latent class analysis^15^) or to a few higher-order dimensions (e.g., threat vs deprivation^16^) are unlikely to capture the full heterogeneity of ACE exposure profiles. While these frameworks have been highly influential, more flexible approaches that focus directly on observed patterns of ACE exposure and make minimal assumptions about how adversities should be grouped may provide a useful complement. Third, routine reporting should include simple *heterogeneity diagnostics* such as combinatorial coverage and UpSet plots to summarise observed patterns and guide further analyses. Finally, sensitivity analyses for coding choices are crucial, as different cut-offs can alter realised item combinations.

Our findings support previously reported concerns about the use of ACE scores in clinical practice:^17^ children with the same count can have markedly different ACE exposure profiles. In our data, just 50 higher-prevalence patterns captured about two-thirds of children with ≥2 ACEs, while the remaining third were scattered across hundreds of very rare combinations. Some constellations are therefore relatively common (e.g., parental separation/divorce with economic hardship), around which clinicians and services can plausibly build more standardised pathways, whereas many others are idiosyncratic and may require more tailored responses to the specific co-occurrence pattern.

At the policy level, these findings align with existing concerns about routine ACE screening in health care settings. Current screening approaches based on simple ACE counts appear to have limited predictive accuracy for later health problems, with substantial numbers of both false positives and false negatives.^18^ Our results demonstrate how substantial item-level heterogeneity under the same ACE score may contribute to this limited discrimination.

## Limitations

This study has some limitations. First, although our data covered expanded ACE categories, including economic hardship, neighbourhood violence, and discrimination,^19^ it did not include some of the original ACE Study items, such as child abuse and neglect.^1^ Second, our ACE measures were based on caregiver report, and the recency of reported events may vary by the child’s age; there are well-documented discrepancies between prospective and retrospective measures and between informants.^20,21^ Taken together, the available ACE items and the method of reporting are likely to influence the observed co-occurrence patterns.

## CONCLUSIONS

ACE scores mask substantial heterogeneity across the 2-, 3-, and ≥4-ACE exposure categories, risking misleading inference about who is at highest risk. A simple quantitative metric – comparing observed vs. possible ACE item combinations (CC) – and visualisation techniques – such as UpSet plots – make this heterogeneity explicit. However, these tools cannot tell us much about variation in effect sizes and should therefore prompt, not replace, pattern-specific analyses of outcomes. We recommend routine reporting of CC and UpSet plots, and, more broadly, suggest that future work consider strategies to address the limitations of cumulative risk and other ACE measurement approaches.

## Data Availability

The 2023 National Survey of Children′s Health (NSCH) datasets and codebooks are publicly available from the US Census Bureau website.

https://www.census.gov/programs-surveys/nsch/data/datasets/nsch2023.html

## Acknowledgements

This study uses data from the 2023 National Survey of Children’s Health (NSCH), funded and directed by the Health Resources and Services Administration (HRSA) Maternal and Child Health Bureau (MCHB), and conducted by the US Census Bureau. Bruna Cardone Pedroso Elorza and Andreas Bauer had full access to all the data in the study and take responsibility for the integrity of the data and the accuracy of the data analysis.

## Data Sharing Statement

The 2023 National Survey of Children’s Health (NSCH) datasets and codebooks are publicly available from the US Census Bureau website.

## Disclaimer

The analyses, interpretations, and conclusions are those of the authors and do not necessarily reflect the views of the Health Resources and Services Administration (HRSA) Maternal and Child Health Bureau (MCHB) or the US Census Bureau.

## Author contributions

**Bruna Cardone Pedroso Elorza:** Conceptualisation, Methodology, Formal analysis, Data Curation, Writing – Original Draft, Writing – Review & Editing, Visualisation, Funding acquisition. **Alicia Matijasevich:** Conceptualisation, Writing – Review & Editing. **Maria Fernanda Tourinho Peres:** Conceptualisation, Writing – Review & Editing. **Andreas Bauer:** Conceptualisation, Methodology, Formal analysis, Data Curation, Writing – Original Draft, Writing – Review & Editing, Visualisation, Supervision, Project administration, Funding acquisition.

## Footnotes

a A PubMed search conducted in December 2025, using the search terms “adverse childhood experiences”[Title/Abstract]) AND (cumulative[Title/Abstract] OR score[Title/Abstract]), yielded almost 1,000 publications from the past five years (2020-2025).

b The 2024 NSCH was not publicly available at the time this study was conducted.

