## Supplementary material for "Quantifying and visualising heterogeneity in cumulative adverse childhood experiences scores": R code: R code_public version.R

```
#################
# Load packages #
#################
library(haven)
library(dplyr)
library(ggplot2)
library(ComplexUpset)
library(naniar)
library(sjmisc)

#####################
# Prepare data file #
#####################
# Load data
df <- haven::read_dta("nsch_2023e_topical.dta")

# Select ACE items and weight variable
dat0 <- df %>%
  dplyr::select(ace1, ace3, ace4, ace5, ace6, ace7, ace8, ace9, ace10, ace11, fwc)

# Recode ACE items to 0/1  
dat1 <- dat0 %>%
  dplyr::mutate(
    # Recode 'economic hardship' (for main analysis)
    ace1 = dplyr::case_when(
      ace1 %in% c(3, 4) ~ 1L,    # somewhat/very often
      ace1 %in% c(1, 2) ~ 0L,    # never/rarely
      TRUE ~ NA_integer_
    ),
    # All other ACEs
    dplyr::across(
      c(ace3, ace4, ace5, ace6, ace7, ace8, ace9, ace10, ace11),
      ~ dplyr::case_when(. == 1 ~ 1L,
                         . == 2 ~ 0L,
                         TRUE   ~ NA_integer_)
    )
  )

# Rename ACE columns 
dat2 <- dat1 %>%
  dplyr::rename(
    `Economic hardship`                           = ace1,
    `Parental divorce/separation`                 = ace3,
    `Household mental illness`                    = ace8,
    `Household substance use`                     = ace9,
    `Parental incarceration`                      = ace5,
    `Domestic violence`                           = ace6,
    `Racial discrimination`                       = ace10,
    `Neighbourhood violence`                      = ace7,
    `Health- or disability-based discrimination`  = ace11,
    `Parental death`                              = ace4
  )

# Define ordered vector of ACE set names for figures
sets <- c(
  "Economic hardship",
  "Parental divorce/separation",
  "Household mental illness",
  "Household substance use",
  "Parental incarceration",
  "Domestic violence",
  "Racial discrimination",
  "Neighbourhood violence",
  "Health- or disability-based discrimination",
  "Parental death"
)

# Restrict to complete ACE data and create ACE score
dat_cc <- dat2 %>%
  dplyr::filter(dplyr::if_all(all_of(sets), ~ !is.na(.))) %>%
  dplyr::mutate(
    ace_count = rowSums(dplyr::across(all_of(sets))),
    ace_cat   = dplyr::case_when(
      ace_count %in% 0:3 ~ as.character(ace_count),
      ace_count >= 4     ~ "4+"
    )
  )

#############
# Bar chart #
#############
# Possible patterns within each ACE exposure category
possible_2  <- choose(10, 2)                 # 45
possible_3  <- choose(10, 3)                 # 120
possible_4p <- 2^10 - sum(choose(10, 0:3))   # 848

# Observed patterns within each ACE exposure category
obs_2  <- dat_cc %>% dplyr::filter(ace_count == 2)  %>%
  dplyr::distinct(dplyr::across(all_of(sets))) %>% nrow() # 45
obs_3  <- dat_cc %>% dplyr::filter(ace_count == 3)  %>%
  dplyr::distinct(dplyr::across(all_of(sets))) %>% nrow() # 110
obs_4p <- dat_cc %>% dplyr::filter(ace_count >= 4) %>%
  dplyr::distinct(dplyr::across(all_of(sets))) %>% nrow() # 427

# Create data frames for plotting 
totals_df <- tibble::tibble(
  Category = factor(c("2 ACEs","3 ACEs","≥4 ACEs"),
                    levels = c("2 ACEs","3 ACEs","≥4 ACEs")),
  Total = c(possible_2, possible_3, possible_4p)
)
observed_df <- tibble::tibble(
  Category = factor(c("2 ACEs","3 ACEs","≥4 ACEs"),
                    levels = c("2 ACEs","3 ACEs","≥4 ACEs")),
  Observed = c(obs_2, obs_3, obs_4p)
)

# Define y-axis breaks 
y_max    <- max(totals_df$Total, na.rm = TRUE)
y_breaks <- seq(0, ceiling(y_max/100)*100, by = 100)

# Define colours 
col_dark <- "#004C99"  # observed 
col_lite <- "#BFD7EA"  # possible

# Define legend position 
panel_top <- max(y_breaks, na.rm = TRUE) * 1.05   
legend_y  <- min(max(690 / panel_top, 0.05), 0.95)
legend_x  <- 0.35                                  

ggplot() +
  # Number of possible patterns
  geom_col(
    data = totals_df,
    aes(x = Category, y = Total, fill = "# of possible patterns"),
    width = 0.75,
    show.legend = TRUE,
    alpha = 0.60
  ) +
  # Number of observed patterns 
  geom_col(
    data = observed_df,
    aes(x = Category, y = Observed, fill = "# of observed patterns"),
    width = 0.75,
    show.legend = TRUE
  ) +
  scale_fill_manual(
    values = c(
      "# of observed patterns"     = col_dark,
      "# of possible patterns" = col_lite
    ),
    breaks = c("# of observed patterns", "# of possible patterns"),
    name   = NULL,
    drop   = FALSE
  ) +
  scale_y_continuous(breaks = y_breaks, expand = expansion(mult = c(0, 0.05))) +
  labs(x = NULL, y = NULL) +   
  theme_minimal(base_size = 13) +
  theme(
    legend.position         = "inside",
    legend.position.inside  = c(legend_x, legend_y),
    legend.direction        = "vertical",
    legend.justification    = c(0.5, 0.5),
    legend.background       = element_blank(),
    legend.key              = element_blank(),
    panel.grid.minor        = element_blank(),
    panel.grid.major.x      = element_blank(),
    panel.grid.major.y = element_line(colour = "grey85", linewidth = 0.5)
  ) +
  guides(fill = guide_legend(ncol = 1, override.aes = list(alpha = c(1, 0.60))))

##############
# UpSet plot #
##############
# Limit data to 2+ ACEs and keep only ACE indicators 
dat_2plus <- dat_cc %>%
  dplyr::filter(ace_count >= 2) %>%
  dplyr::select(dplyr::all_of(sets)) # n = 7,897

# Limit to intersections of n >= 30 
comb_labels <- apply(dat_2plus[, sets, drop = FALSE], 1L, function(x) {
  on <- sets[which(x == 1L)]
  if (length(on) == 0L) "" else paste(on, collapse = " & ")
})
tab        <- sort(table(comb_labels), decreasing = TRUE)
keep_names <- names(tab)[tab >= 30]

# Show these intersections as table
upset_counts <- tibble::tibble(
  pattern = keep_names,
  n       = as.integer(tab[keep_names])
)
print(upset_counts, n = 50) # UpSet plot intersections as table

n_2plus <- sum(tab)
n_2plus # total number of children with >= 2 ACEs
n_displayed_30 <- sum(tab[keep_names])
n_displayed_30 # number of children with patterns with n >= 30
n_not_displayed_30 <- n_2plus - n_displayed_30
n_not_displayed_30 # number of children with patterns with n < 30

# Define colours by exposure category 
col_k2  <- "#D95F02"   # 2 ACEs
col_k3  <- "#1B9E77"   # 3 ACEs
col_k4p <- "#7570B3"   # 4+ ACEs

# Build query list to colour intersections by exposure group 
queries_colour <- lapply(keep_names, function(lbl) {
  items <- strsplit(lbl, " & ", fixed = TRUE)[[1]]
  k     <- length(items)
  col   <- if (k == 2) col_k2 else if (k == 3) col_k3 else col_k4p
  ComplexUpset::upset_query(
    intersect       = items,
    only_components = c("Intersection size","intersections_matrix"),
    fill            = col,
    color           = col
  )
})

# NOTE: If you only need the plot (with minimal customisation),
# you can skip the previous steps and use the ComplexUpset's built-in functions:
# - min_degree = 2 (restrict to intersections with 2+ ACEs)
# - min_size = 30 (limit to intersections with n >= 30)

# Create UpSet plot (unweighted)
up_overall_unw <- ComplexUpset::upset(
  dat_2plus,
  intersect             = sets,
  encode_sets           = FALSE,                 
  min_size              = 30,
  sort_intersections    = "descending",
  sort_intersections_by = "cardinality",
  set_sizes             = FALSE,
  name                  = " ",
  queries               = queries_colour,
  # Modify position of top bar panel and matrix (optional)
  themes = ComplexUpset::upset_modify_themes(list(
    "Intersection size" = ggplot2::theme(
      plot.margin = ggplot2::margin(t = 0, r = 0, b = -6, l = 0)  
    ),
    "intersections_matrix" = ggplot2::theme(
      plot.margin = ggplot2::margin(t = -6, r = 0, b = 0, l = 0),  
      axis.text.y  = ggplot2::element_text(size = 11.5)
    )
  )),
  # Modify matrix (optional)
  matrix = ComplexUpset::intersection_matrix(
    geom = ggplot2::geom_point(
      shape = 19,         
      size  = 3,        
      alpha = 1        
    ),
    segment = ggplot2::geom_segment(
      linewidth = 1.05,   
      alpha     = 1,
      lineend   = "round"
    ),
    outline_color = list(active = NA, inactive = NA)   
  ),
  # Modify top bars (optional)
  base_annotations = list(
    "Intersection size" =
      ComplexUpset::intersection_size(
        counts = TRUE,
        width = 0.80,
        text   = list(size = 2.8, vjust = -0.35, check_overlap = TRUE)
      ) +
      ggplot2::expand_limits(y = 700) +
      ggplot2::scale_y_continuous(
        breaks = seq(0, 700, by = 100),
        minor_breaks = NULL,
        expand = ggplot2::expansion(mult = c(0, 0.05))
      ) +
      ggplot2::theme(
        panel.grid.major.y = ggplot2::element_line(colour = "grey85", linewidth = 0.4),
        panel.grid.minor.y = ggplot2::element_blank(),
        panel.grid.major.x = ggplot2::element_blank()
      )
  )
)
up_overall_unw

################################################
# Weighted estimates for selected ACE patterns #
################################################
# 2 ACEs
w_divorce_hardship <- dat_cc %>%
  filter(`Parental divorce/separation` == 1,
         `Household substance use` == 0,
         `Household mental illness` == 0,
         `Economic hardship` == 1,
         `Parental incarceration` == 0,
         `Domestic violence` == 0,
         `Neighbourhood violence` == 0,
         `Racial discrimination` == 0,
         `Health- or disability-based discrimination` == 0,
         `Parental death` == 0) %>%
  summarise(weighted_exclusive_N = sum(fwc, na.rm = TRUE)) %>%
  pull()
w_divorce_hardship

w_divorce_subuse <- dat_cc %>%
  filter(`Parental divorce/separation` == 1,
         `Household substance use` == 1,
         `Household mental illness` == 0,
         `Economic hardship` == 0,
         `Parental incarceration` == 0,
         `Domestic violence` == 0,
         `Neighbourhood violence` == 0,
         `Racial discrimination` == 0,
         `Health- or disability-based discrimination` == 0,
         `Parental death` == 0) %>%
  summarise(weighted_exclusive_N = sum(fwc, na.rm = TRUE)) %>%
  pull()
w_divorce_subuse

w_divorce_mentill <- dat_cc %>%
  filter(`Parental divorce/separation` == 1,
         `Household substance use` == 0,
         `Household mental illness` == 1,
         `Economic hardship` == 0,
         `Parental incarceration` == 0,
         `Domestic violence` == 0,
         `Neighbourhood violence` == 0,
         `Racial discrimination` == 0,
         `Health- or disability-based discrimination` == 0,
         `Parental death` == 0) %>%
  summarise(weighted_exclusive_N = sum(fwc, na.rm = TRUE)) %>%
  pull()
w_divorce_mentill
# 3 ACEs
w_divorce_subuse_mentill <- dat_cc %>%
  filter(`Parental divorce/separation` == 1,
         `Household substance use` == 1,
         `Household mental illness` == 1,
         `Economic hardship` == 0,
         `Parental incarceration` == 0,
         `Domestic violence` == 0,
         `Neighbourhood violence` == 0,
         `Racial discrimination` == 0,
         `Health- or disability-based discrimination` == 0,
         `Parental death` == 0) %>%
  summarise(weighted_exclusive_N = sum(fwc, na.rm = TRUE)) %>%
  pull()
w_divorce_subuse_mentill

w_divorce_subuse_parinc <- dat_cc %>%
  filter(`Parental divorce/separation` == 1,
         `Household substance use` == 1,
         `Household mental illness` == 0,
         `Economic hardship` == 0,
         `Parental incarceration` == 1,
         `Domestic violence` == 0,
         `Neighbourhood violence` == 0,
         `Racial discrimination` == 0,
         `Health- or disability-based discrimination` == 0,
         `Parental death` == 0) %>%
  summarise(weighted_exclusive_N = sum(fwc, na.rm = TRUE)) %>%
  pull()
w_divorce_subuse_parinc

w_divorce_mentill_hardship <- dat_cc %>%
  filter(`Parental divorce/separation` == 1,
         `Household substance use` == 0,
         `Household mental illness` == 1,
         `Economic hardship` == 1,
         `Parental incarceration` == 0,
         `Domestic violence` == 0,
         `Neighbourhood violence` == 0,
         `Racial discrimination` == 0,
         `Health- or disability-based discrimination` == 0,
         `Parental death` == 0) %>%
  summarise(weighted_exclusive_N = sum(fwc, na.rm = TRUE)) %>%
  pull()
w_divorce_mentill_hardship
# 4+ ACEs
w_divorce_subuse_mentill_parinc_DV_NV <- dat_cc %>%
  filter(`Parental divorce/separation` == 1,
         `Household substance use` == 1,
         `Household mental illness` == 1,
         `Economic hardship` == 0,
         `Parental incarceration` == 1,
         `Domestic violence` == 1,
         `Neighbourhood violence` == 1,
         `Racial discrimination` == 0,
         `Health- or disability-based discrimination` == 0,
         `Parental death` == 0) %>%
  summarise(weighted_exclusive_N = sum(fwc, na.rm = TRUE)) %>%
  pull()
w_divorce_subuse_mentill_parinc_DV_NV

w_divorce_subuse_mentill_parinc_DV <- dat_cc %>%
  filter(`Parental divorce/separation` == 1,
         `Household substance use` == 1,
         `Household mental illness` == 1,
         `Economic hardship` == 0,
         `Parental incarceration` == 1,
         `Domestic violence` == 1,
         `Neighbourhood violence` == 0,
         `Racial discrimination` == 0,
         `Health- or disability-based discrimination` == 0,
         `Parental death` == 0) %>%
  summarise(weighted_exclusive_N = sum(fwc, na.rm = TRUE)) %>%
  pull()
w_divorce_subuse_mentill_parinc_DV

w_divorce_subuse_mentill_DV <- dat_cc %>%
  filter(`Parental divorce/separation` == 1,
         `Household substance use` == 1,
         `Household mental illness` == 1,
         `Economic hardship` == 0,
         `Parental incarceration` == 0,
         `Domestic violence` == 1,
         `Neighbourhood violence` == 0,
         `Racial discrimination` == 0,
         `Health- or disability-based discrimination` == 0,
         `Parental death` == 0) %>%
  summarise(weighted_exclusive_N = sum(fwc, na.rm = TRUE)) %>%
  pull()
w_divorce_subuse_mentill_DV

###########################
# Missing data UpSet plot #
###########################
miss_df <- dat1[, c("ace1","ace3","ace8","ace9","ace5","ace6","ace10","ace7","ace11","ace4")]
names(miss_df) <- c(
  "Economic hardship",
  "Parental divorce/separation",
  "Household mental illness",
  "Household substance use",
  "Parental incarceration",
  "Domestic violence",
  "Racial discrimination",
  "Neighbourhood violence",
  "Health- or disability-based discrimination",
  "Parental death"
)
miss_df$`Any ACE missing` <- rowSums(is.na(miss_df)) > 0
for (v in names(miss_df)) print(sjmisc::frq(miss_df[[v]], show.na = TRUE))
miss_df_upset <- miss_df[, -11]
naniar::gg_miss_upset(miss_df_upset,
                      nsets = 10,
                      nintersects = 12) # n >= 30

##################################
# Sensitivity analysis re-coding #
##################################
# Run and then re-run code from above
dat1 <- dat0 %>%
  dplyr::mutate(
    # Recode 'economic hardship' 
    ace1 = dplyr::case_when(
      ace1 %in% c(2, 3, 4) ~ 1L,    # rarely/somewhat/very often
      ace1 == 1            ~ 0L,    # never
      TRUE ~ NA_integer_
    ),
    # All other ACEs
    dplyr::across(
      c(ace3, ace4, ace5, ace6, ace7, ace8, ace9, ace10, ace11),
      ~ dplyr::case_when(. == 1 ~ 1L,
                         . == 2 ~ 0L,
                         TRUE   ~ NA_integer_)
    )
  )
```
